# Proprietary and Open-Source Large Language Models on the Korean Pharmacist Licensing Examination: A Comparative Benchmarking Study

**DOI:** 10.1101/2025.04.15.25325584

**Authors:** David Hyunyoo Jang, Juyong Lee

## Abstract

**Background:** Large language models (LLMs) have shown remarkable advancements in natural language processing, with increasing interest in their ability to handle tasks requiring expert-level knowledge. While previous studies have evaluated specific LLM models on pharmacist licensing examinations, comprehensive benchmarking across diverse model architectures, sizes, and generations remains limited. This study addresses this gap by systematically evaluating LLM capabilities on the Korean Pharmacist Licensing Examination (KPLE), a high-stakes professional certification test.

**Methods:** We conducted a comprehensive benchmark of 27 LLMs, spanning proprietary models (GPT, Claude, Gemini, PaLM series) and open-source models across three size categories (small: 4-10B, medium: 14-35B, large: 70-104B parameters), using both original Korean and English-translated KPLE examinations from 2019 to 2024. Models were evaluated using accuracy-based and score-based metrics, with systematic analysis of subject-specific performance, temporal progression, cross-linguistic capabilities, and item-level difficulty patterns.

**Results:** Seven models achieved passing scores across all six examination years in both languages, demonstrating substantial progress in LLM capabilities. The top-performing model, Claude 3.5 Sonnet, ranked in the top 12% of human examinees. Temporal analysis revealed rapid improvement, particularly among open-source models, with performance gaps narrowing considerably over the 12-month study period. Parameter size correlated with performance following a logarithmic relationship, though recent architectural innovations enabled smaller models to outperform larger predecessors. Cross-linguistic evaluation showed reduced performance disparities in newer models. Subject-level analysis identified consistent strengths in memorization-intensive topics (Biopharmacy) and weaknesses in domains requiring complex calculations (Physical Pharmacy, Pharmaceutical Analysis) and region-specific knowledge (Medical Health Legislation, Pharmaceutical Quality Science).

**Conclusion:** This comprehensive benchmarking study demonstrates that current LLMs can successfully pass the KPLE, with capabilities spanning diverse model architectures and sizes. Performance improvements are driven by multiple factors including parameter scaling, architectural innovations, enhanced multilingual training data, and fine-tuning strategies. Models excel in memorization and language comprehension but show limitations in complex reasoning and nation-specific knowledge domains. These findings highlight opportunities for targeted improvement through domain-specific fine-tuning and specialized training. While LLMs cannot substitute for human pharmacists, they show promise as complementary tools for education, decision support, and administrative tasks. Future development should focus on addressing identified weaknesses while leveraging the distinct advantages of both proprietary and open-source approaches to ensure safe and effective pharmaceutical applications.

## 1 Background

In recent years, the advancement of large language models (LLMs) has marked significant progress in the field of artificial intelligence, particularly in natural language processing (NLP). The development of models such as OpenAI’s GPT-3, released as the foundation of ChatGPT in November 2022, has sparked considerable interest and investment in LLM technology [1]. The subsequent release of GPT-4 further propelled interest in LLMs due to its superior performance in a variety of tasks [2]. Concurrently, other major technology companies have introduced their own LLMs, such as Google’s PaLM series [3, 4] (later evolving into Gemini [5, 6]) and Meta’s Llama series [7, 8, 9], further intensifying research and competition in this domain. These developments have accelerated research and competition within the LLM landscape.

Recent LLMs have demonstrated capabilities approaching or surpassing human-level performance across various language understanding and generation tasks [2]. This has led to a growing interest in the potential of LLMs to perform in domains requiring specialized knowledge that have traditionally been reserved for human experts. Notable examples include GPT-4 achieving scores above the passing threshold on the US bar examination and medical licensing examination [2, 10, 11]. Similarly, Google’s Med-PaLM, trained primarily on medical domain-specific information, highlights the growing trend of leveraging LLMs in specialized fields [12].

Recent research has examined LLM performance on pharmacist licensing examinations across various countries, revealing both capabilities and consistent limitations. Studies on the Japanese examination found GPT-4 achieved 78% accuracy on text-based questions but struggled with diagram-dependent content (17% exclusion rate) [13, 14]. The Taiwan examination revealed GPT-3.5 failed to pass (54% accuracy), with notable performance differences between Chinese and English versions [15]. In Chinese medical licensing examinations, GPT-4 significantly outperformed GPT-3.5, though both models showed weaknesses in specialized domains [16]. Evaluation on the Korean examination demonstrated GPT-4’s passing capability (86% accuracy), substantially exceeding GPT-3.5 performance (60% accuracy) [17]. Assessment using North American NAPLEX practice questions showed LLMs approaching but not consistently exceeding passing thresholds [18]. These studies collectively identify common patterns: (1) marked performance improvements from GPT-3.5 to GPT-4, (2) superior performance in memorization-based domains compared to calculation-intensive topics, (3) challenges with nation-specific regulatory content, and (4) methodological difficulties handling visual content, where exclusion rates range from 8% to 17% depending on examination structure.

This study builds upon this backdrop to evaluate the performance of LLMs in a specific, high-stakes professional domain: the Korean Pharmacist Licensing Examination (KPLE) [19]. The KPLE is a comprehensive and rigorous examination essential for obtaining a pharmacist license in South Korea, as stipulated by Article 3, Section 2 of the Pharmaceutical Affairs Act. To obtain a pharmacist license, candidates must graduate from an accredited pharmacy program and pass the KPLE, administered by the Korea Health Personnel Licensing Examination Institute (KHPLEI). The examination assesses knowledge across four major subjects: Biopharmacy, Industrial Pharmacy, Clinical & Practical Pharmacy, and Medical Health Legislation, each encompassing a wide array of topics reflective of the pharmacy curriculum.

This study has four primary objectives. First, we aim to benchmark the performance of both proprietary and open-source large language models (LLMs) on the KPLE. To achieve this, we evaluate proprietary models including GPT-4 and Claude 3, alongside open-source counterparts including Llama 3 and Qwen 2. Second, we seek to determine whether current LLMs can successfully pass the KPLE by assessing their proficiency in specialized professional examinations. Third, we analyze the strengths and weaknesses of LLMs in comparison to real examinees, offering insights into areas where these models excel or require improvement. Finally, we explore the implications of LLM performance for the future role of pharmacists, considering how artificial intelligence (AI) integration might enhance or transform professional practices in the field of pharmacy.

Given the structure and requirements of the KPLE, it presents an unique challenge for LLMs in determining whether these models can effectively replicate the specialized knowledge and critical reasoning required to pass such an examination. The choice of the KPLE is significant due to its high standards and the critical nature of the knowledge it tests, making it an ideal tool for exploring the potential of LLMs in professional and academic settings. Our results highlight how training data and methodologies influence LLM performance in tasks that require deep, domain-specific knowledge. By analyzing translated items and culturally adapted content, this study emphasizes the need for diverse datasets in LLM development to ensure robust performance across different languages and cultural contexts.

## 2 Methods

### 2.1 Model Selection

This study utilizes both proprietary and open-source LLMs to benchmark their performance on the Korean Pharmacist Licensing Examination (KPLE). The selection encompasses a diverse range of models, including text generation models and instruction-tuned models, to comprehensively assess their capabilities in a specialized professional domain. The detailed information on the models used in this study is provided in Table 1.

**Table 1:**
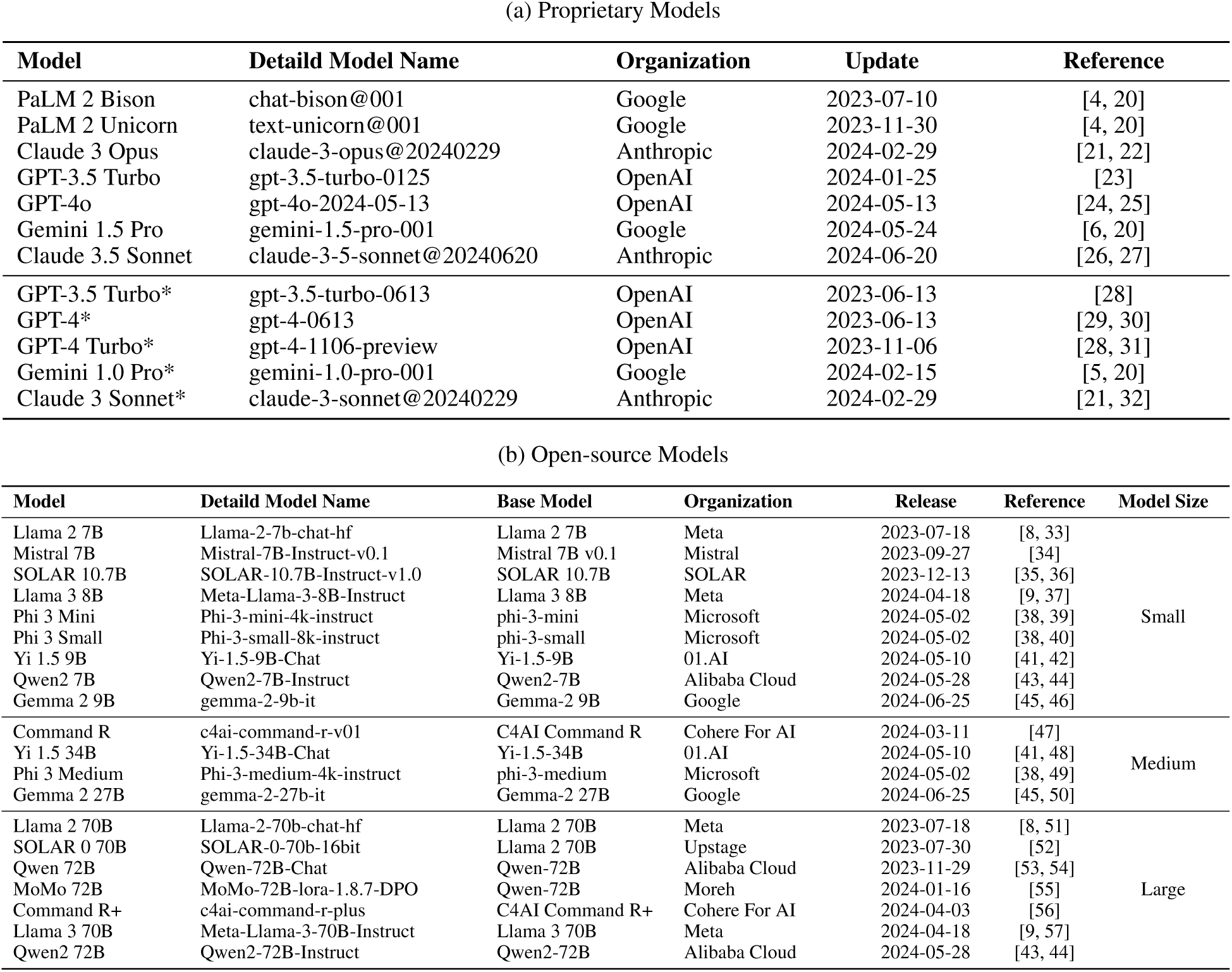
Detailed Information on Selected Models.

**Table 2:**
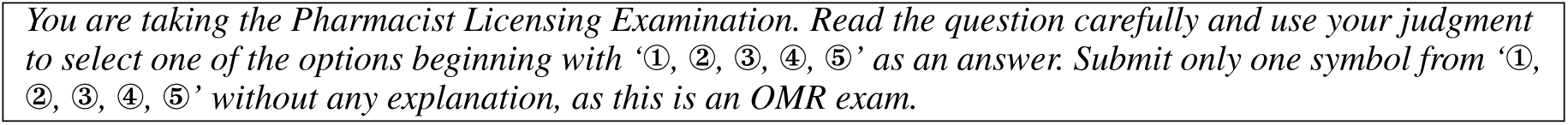
Instruction for Models.

#### 2.1.1 Types of Models

Currently available LLMs can be categorized into two groups: text gereration models and instruction tuned models. Generally, text generation models are pre-trained on extensive corpora of text data and operate on a decoder-only architecture, predicting the next token based on given input tokens. In contrast, instruction-tuned models are fine-tuned for specific tasks such as chat, question-answering, or summarization. These models employ distinct tokens to differentiate user instructions and inputs from the model-generated outputs, enhancing their ability to follow conversational prompts effectively.

Our preliminary tests indicated that text generation models tend to generate new instructions or questions rather than providing direct solutions to problems (Supplementary Table 1). Conversely, instruction-tuned models demonstrated superior performance in solving questions due to their conversational design. Consequently, this study primarily focuses on instruction-tuned models for the KPLE exam, with the exception of PaLM 2 Unicorn, which lacks a chat-optimized version.

Instruction-tuned models typically employ three roles—system, user, and assistant—to maintain dialogue flow and coherence. While specific prompt formats varied slightly depending on the model, the general structure includes a system role providing the direction, an user role with the question, and an assistant role generating responses with specific formats. The prompt format example for Llama 2 70B is presented in Supplementary Table 1. The following instruction was consistently used across all prompts:

#### 2.1.2 Model Categorization

In this study we tested 27 models released between June 2023 and June 2024. These models are divided into two main categories. The first category includes seven proprietary models, including both the latest and legacy versions, to facilitate comparative analysis. The second category consists of 20 open-source models, which are further sub-divided based on their computational resource requirements. Small models can run on a single GPU with 24GB of VRAM. Medium models require multiple GPUs for execution. Large models necessitate six or more GPUs due to their extensive parameter sizes.

#### 2.1.3 Rationale for Model Selection

The chosen models represent a broad spectrum of architectures and training methodologies, allowing for a nuanced analysis of their performance on the KPLE. By including both proprietary and open-source models, we aim to identify performance trends related to model size, architecture advancements, and the impact of fine-tuning on specialized tasks. This diverse selection is critical in benchmarking the state-of-the-art LLMs and understanding their applicability in professional examinations such as the KPLE.

### 2.2 Preparing KPLE Questions and Scoring Results

The KPLE exam consists of 350 questions covering four subjects, as described in Table 3. To pass the exam, candidates must achieve at least a 40% scaled percentage score in each subject and an overall score of 60% or higher are required. Historical data from the 70th to the 75th editions of the exam ranging form year 2019 to 2024, were obtained from the KHPLEI online platform [58] and analyzed accordingly.

**Table 3:** Subjects and Details of KPLE.

| Subject | Identifier | Topic | Number of Questions |
| --- | --- | --- | --- |
| **Biopharmacy** | 1-1 | Biochemistry | 20 |
|  | 1-2 | Microbiology & Immunology | 20 |
|  | 1-3 | Pharmacology | 20 |
|  | 1-4 | Preventive Pharmacy | 20 |
|  | 1-5 | Pathophysiology | 20 |
| **Industrial Pharmacy** | 2-1 | Physical Pharmacy | 18 |
|  | 2-2 | Pharmaceutical & Medicinal Chemistry | 18 |
|  | 2-3 | Pharmaceutical Analysis | 18 |
|  | 2-4 | Pharmaceutics | 18 |
|  | 2-5 | Pharmacognosy | 18 |
| **Clinical & Practical Pharmacy** | 3-1 | Pharmacotherapy | 77 |
|  | 3-2 | Pharmacy Practice | 27 |
|  | 3-3 | Pharmaceutical Quality Science | 18 |
|  | 3-4 | Social Pharmacy | 18 |
| **Medical Health Legislation** | 4 | Enforcement degrees and rules of *Pharmaceutical Affairs Act*, *Narcotics Control Act*, *National Health Promotion Act*, *Framework Act on Health and Medical Services*, *National Health Insurance Act*, *Regional Public Health Act* of the Republic of Korea | 20 |

The questions from the 70th to the 74th editions were extracted from PDF files using Python scripts and formatted into JSON files. Necessary adjustments were made for any missing information or tabular data. The 75th edition was provided as a non-selectable text PDF, requiring the use of GPT-4o’s vision capabilities to recognize and extract the text [24]. Answers were collected in a similar manner and compiled into CSV files for structured analysis.

Exam questions were translated from Korean to English using multiple language models, including GPT-4, Claude 3.5 Sonnet, and DeepL. In two preliminary tests, the questions translated by GPT-4 received higher scores than those translated by DeepL, and the questions translated by Claude 3.5 Sonnet received higher scores than those translated by GPT-4 (Supplementary Table 2). Therefore, among the candidates, the questions translated by Claude 3.5 Sonnet were ultimately used in the study. The majority of this study was primarily conducted using the English-translated version of the exam. Meanwhile the original Korean version was only used for evaluating language-based performance differences. The examples of the KPLE questions are shown in Supplementary Table 3.

To ensure consistency and readability, subject names were introduced with both their full names and unique identifiers for ease of reference in subsequent sections. For instance, the Pharmacotherapy topic within Clinical & Practical Pharmacy was labeled as ‘3-1’.

Furthermore, we categorized the exam questions into two types: knowledge-based items, assessing power of recall and comprehension, and skills-based items, evaluating advanced cognitive abilities such as calculation or interpretation.

### 2.3 Edited Questions

Due to copyright restrictions, all graphic content, including chemical structures, reaction equations, and plots, were excluded. Out of the 2,100 questions analyzed across all exam editions, 160 were edited due to the presence of such visual elements. These questions, even with edited diagrams or charts, were included in grading to ensure consistency and facilitate a comprehensive evaluation process.

The proportion of edited questions varied across subjects. Biopharmacy had 4.5% edited questions. Industrial Pharmacy had 22.0%. Clinical & Practical Pharmacy had 1.7%. Medical Health Legislation had no edited questions. Industrial Pharmacy exhibited the highest exclusion rate due to its reliance on visual elements such as chemical structures and reaction equations. In contrast, Biopharmacy and Clinical & Practical Pharmacy had lower exclusion rates, reflecting their lesser dependence on visual data.

At the topic level, exclusion rates also varied significantly. Among all topics, Pharmaceutical & Medicinal Chemistry [2–2], which involves chemical structures, exhibited the highest exclusion rate of 81.5% of its questions being edited. Other topics with high exclusive rate included Biochemistry [1–1], Physical Pharmacy [2–1], and Pharmaceutical Analysis [2–3], with exclusion rates of 17.5%, 10.2%, and 13.0%, respectively. The edited questions in these areas often included reaction equations or complex plots. Most topics retained a sufficient number of unedited questions (at least 94 per category); however, topic 2-2 had only 20 unedited questions across all six years, which could impact the statistical significance of analyses involving this category.

Similar exclusion approaches have been reported in other pharmacist examination studies. Kunitsu [13] excluded 17.4% (60/344) and 16.5% (57/345) of questions from the Japanese pharmacist examination due to diagram requirements. Our overall exclusion rate of 7.6% (160/2,100) reflects the KPLE’s particular design characteristics, though subject-specific rates vary considerably. The handling of visual content represents a significant limitation for current text-based LLMs, though recent multimodal models show promise in addressing this challenge [24]. As Kunitsu [13] noted, questions requiring diagram interpretation are essential to pharmacists’ ability to infer drug characteristics from structural formulas or predict chemical changes, highlighting an important area for future LLM development.

### 2.4 Analysis and Evaluation Methods

#### 2.4.1 Score-based anaylysis Comparison

To evaluate model performance, we administered the exam to 27 models and compared their scores. The primary analysis utilized percentage-based scores aligned with the official scoring structure of the KPLE exam. Pass/fail determinations followed the official criteria, requiring an average score of at least 40% in each subject and an overall average of 60% or higher to be considered passing.

To enable a comparative analysis with human examinees, we also calculated percentile scores. The percentile scores obtained were normalized using the six-year historical average (2019–2024) and standard deviation of the KPLE exam, providing a standardized measure of relative performance. By employing both percentage-based absolute scores and percentile-based relative rankings, we aimed to offer a more comprehensive assessment of model performance.

To analyze the performance of LLMs over time, we examined a subset of 12 models, that achieved top rankings on the LLM leaderboard at the time of their release [59, 60]. These models included Claude 3.5 Sonnet, GPT-4o, Llama 3 70B, Claude 3 Opus, PaLM 2 Unicorn, Gemini 1.5 Pro, MoMo 72B, Qwen 72B, GPT-3.5 Turbo, PaLM 2 Bison, SOLAR 0 70B, and Llama 2 70B. For comparison, we also included legacy models such as GPT-4 Turbo, GPT-4, Claude 3 Sonnet, GPT-3.5 Turbo, and Gemini 1.0 Pro.

For a more detailed subject-level analysis, we grouped 19 models into four categories: high-performing proprietary models, high-performing large models, medium-sized models, and small models. The proprietary models included Claude 3.5 Sonnet, GPT-4o, Claude 3 Opus, PaLM 2 Unicorn, and Gemini 1.5 Pro. The large models consisted of Qwen2 72B, Llama 3 70B, MoMo 72B, Command R+ (104B), and Qwen 72B. The medium-sized models analyzed were Gemma 2 27B, Yi 1.5 34B, Phi 3 Medium (14B), and Command R (35B), whereas the small models included Qwen2 7B, Llama 3 8B, Phi 3 Small (7B), Gemma 2 9B, and Phi 3 Mini (4B). By comparing these different model categories, we aimed to identify patterns in performance across various model sizes and architectures, providing deeper insights into the strengths and weaknesses of different LLMs at the subject level.

#### 2.4.2 Accuracy-based analysis

To conduct an item-wise analysis, we compared the distribution of item-level accuracy across different subjects for the entire model set, the high-scoring group, and the low-scoring group. The accuracy of each item was defined as the percentage of models that correctly answered the question within a given group. The error rate calculated as 100 minus the accuracy. For this analysis, three outlier models—Llama 2 7B, Llama 2 70B, and Mistral 7B—were excluded, as they consistently failed to pass both the translated and original versions of the KPLE exam. The high-scoring and low-scoring groups were determined by selecting the top 11 and bottom 11 models, respectively. This selection ensured consistency in the subsequent difficulty analysis by maintaining a balanced distribution of performance levels.

To assess question difficulty, we analyzed the percentage of incorrect responses among models in both the high-scoring and low-scoring groups. Questions with higher incorrect response rates were considered more difficult. For both groups, the 11 selected models were used to categorize difficulty levels. Based on the distribution of incorrect answers, questions that zero to three models answered incorrectly were considered ‘Easy’, questions that four to seven models answered incorrectly were considered ‘Medium’, and questions that eight to eleven models answered incorrectly were considered ‘Hard’.

To compare difficulty patterns between the two groups, we calculated the difference in the number of correct responses between high-scoring and low-scoring groups for each item. Items where the difference was four or more models were categorized as either ‘Low>High’ (items more difficult for the low-scoring group) or ‘Low<High’ (items more difficult for the high-scoring group). Items with differences less than four were categorized as ‘LowHigh’ (similar difficulty across both groups). A high proportion of LowHigh items would indicate similar difficulty trends between the groups.

Additionally, we compared these results with human difficulty assessments of KPLE provided by the KHPLEI [61]. Since analysis data from KHPLEI did not include topic-level difficulty analysis, we conducted comparisons at the subject level to examine whether LLMs and human examinees exhibited similar patterns in subject-specific difficulty.

#### 2.4.3 Computational Environment

The open source models were tested on a server equipped with eight NVIDIA RTX A6000 Ada GPUs. The Huggingface Transformers library [62], integrated with FlashAttention-2 [63], was employed for faster inference. The proprietary models were accessed via each model’s Python APIs,

For deterministic results, the open source models were set to non-sampling modes, and the proprietary models utilized zero sampling temperature settings. Preliminary testing revealed that GPT models accessed through OpenAI APIs exhibited non-deterministic behavior even with temperature set to zero. However, the variability was primarily observed in response formatting rather than substantive answer changes—for instance, a correct answer of option 5 might be formatted as ‘⑤’, ‘5’, or ‘⑤ (option text)’, despite of the instruction. Thus, we conducted single runs for all models to make item-level analysis consistent. We acknowledge that answers may vary across runs for questions where models lack confidence. No selective reporting of favorable results was performed; all reported scores represent single-run outcomes.

#### 2.4.4 Software and Libraries

For open-source models, we used Transformers v4.46.2 (https://github.com/huggingface/transformers) and FlashAttention v2.5.9 (https://github.com/Dao-AILab/flash-attention).

For proprietary models, the GPT series were accessed via the OpenAI Python API v1.35.7 (https://github.com/ openai/openai-python). The PaLM, Gemini, and Claude series were accessed using the Vertex AI SDK for Python v1.57.0 (https://github.com/googleapis/python-aiplatform).

For data analysis and visualization, we used Python v3.13.2 (https://www.python.org/), along with the following libraries: NumPy v1.26.3 (https://github.com/numpy/numpy), pandas v2.2.3 (https://github.com/pandas-dev/pandas), Matplotlib v3.10.1 (https://github.com/matplotlib/matplotlib), and seaborn v0.13.2 (https://github.com/mwaskom/seaborn). Microsoft Excel (Microsoft 365 Subscription) was also used for data handling.

## 3 Results

### 3.1 Overall Score Analysis

A total of seven models passed both the original Korean and English-translated versions of the exam across all six years (Figure 1c). Among these models, two were large open-source models (Llama 3 70B and Qwen2 72B), and the remaining five were proprietary models (Claude 3 Opus, GPT-4o, Claude 3 Opus, PaLM 2 Unicorn, Gemini 1.5 Pro).

**Figure 1:**
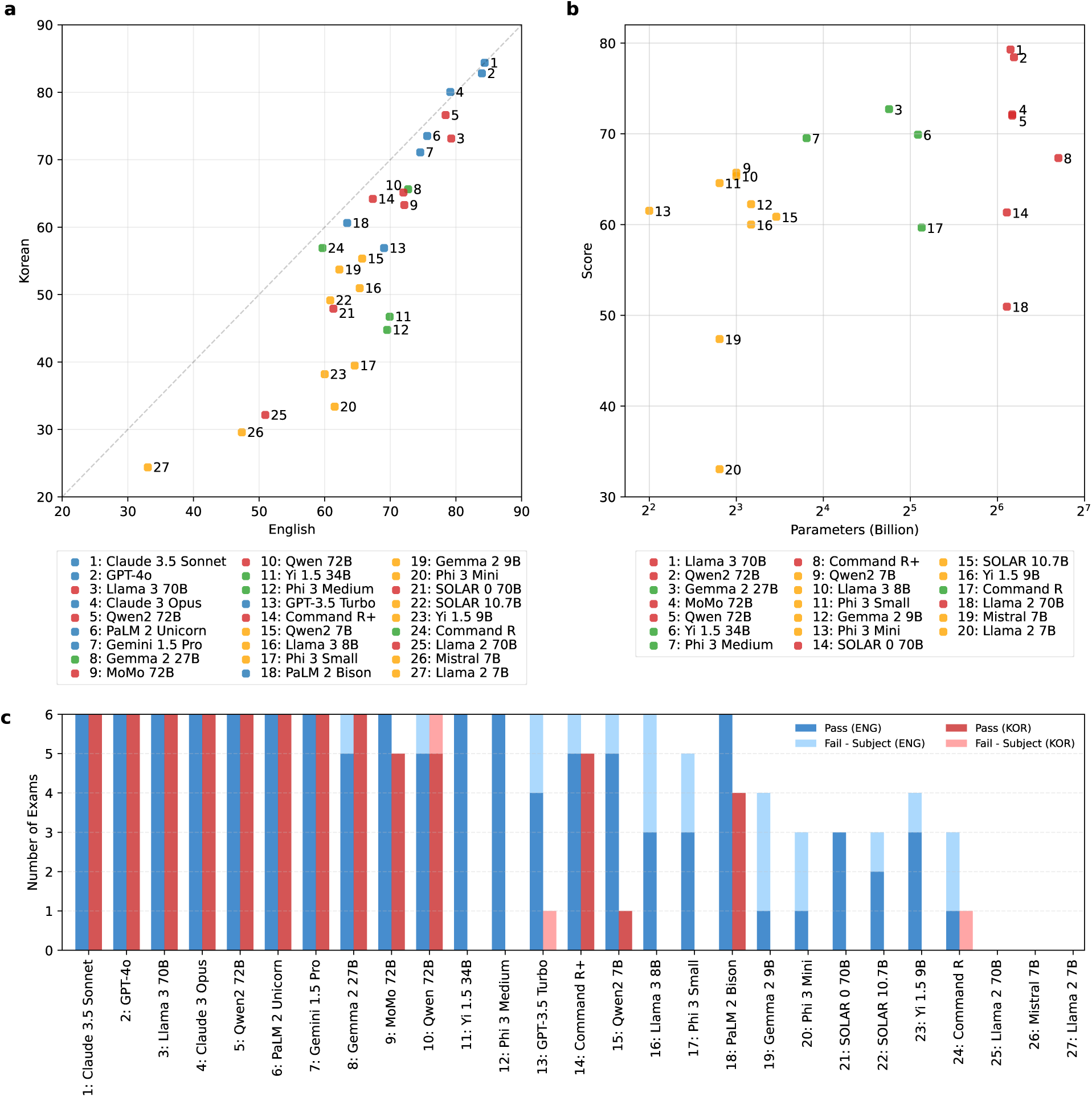
Overall model performance on the KPLE exam. (a) A comparison of scores between English-translated exams (x-axis) and original Korean (y-axis). Each point represents a model; points below the diagonal indicate bet-ter performance in English, while points above indicate better performance in Korean. Models are color-coded by category: proprietary (blue), open-source large (red), medium (green), and small (yellow). (b) Relationship between parameter size (in billions, log scale on x-axis) and model performance scores (y-axis), color-coded as in (a). (c) Stacked bar chart showing the number of passed exams (darker bars) and subject-specific failures (lighter bars, exams that passed 60% overall but failed to meet 40% in at least one subject) for Korean (red) and English (blue) versions, ordered by English exam performance.

Analysis of failure patterns across 162 tests per language (6 years × 27 models) revealed distinct criteria contributing to examination failures. For the Korean version, 91 failures resulted from overall scores below 60%, while only 3 failures occurred due to subject-specific scores below 40% despite meeting the overall threshold. In contrast, the English version showed 35 overall failures and 20 subject-specific failures. Notably, all subject-specific failures were attributable solely to Medical Health Legislation [4], reflecting this subject’s particular challenge for LLMs as discussed in subsequent sections.

Overall, proprietary models achieved higher scores compared to open-source models (Figure 1a). Among proprietary models, Claude 3.5 Sonnet demonstrated the highest performance, scoring 84.4 on both the original Korean and the English-translated exams, successfully passing all six years. Among open-source models, the large model Llama 3 70B achieved the highest score of 79.3 on the English-translated exam, while Qwen 2 72B achieved the highest score of 76.6 on the original Korean exam, with both models passing all tests. In the medium-sized category, Gemma 2 27B scored highest, achieving 65.6 on the Korean version and 72.7 on the English version; notably, it passed all Korean exams but failed one English exam. Among small models, Qwen 15B had the highest performance, scoring 55.3 on the Korean exam and 65.7 on the English exam, passing five of the English exams and one Korean exam.

Models with larger parameter sizes generally obtained higher scores (Figure 1b). However, some recent models with smaller parameter sizes surpassed older models with larger parameter sizes. For example, Gemma 2 27B outperformed all large models except Llama 3 70B and Qwen2 72B, successfully passing five years of English exams. Similarly, Phi 3 Medium successfully passed all six years of English exams, despite having slightly lower scores than Gemma 2 27B and only 14 billion parameters,. In general, among models with comparable parameter sizes, the highest scores tended to show a logarithmic relationship with parameter size.

In most cases, the models achieved higher scores on the English-translated exams compared to the original Korean versions, leading to a greater number of passed exams (Figure 1a & 1b). Models with higher overall performance exhibited smaller discrepancies between their scores on Korean and English exams. However, certain models, such as Yi 1.5 34B and Phi 3 Medium, exhibited extreme disparities, passing all six English exams but failing to pass any of the original Korean exams.

### 3.2 Model Improvement Trends

Both proprietary and open-source models showed performance improvements over time (Figure 2). Among proprietary models, GPT-4 (June 2023) already achieved a score corresponding to the top 25.5% of human examinees (percentile score 74.5) and passed all English-translated exams. Claude 3.5 Sonnet, released in June 2024, improved further, ranking at the top 12.0% of examinees (percentile score 88.0). Over the roughly 12-month interval between GPT-4 and Claude 3.5 Sonnet, raw scores increased by 5.2 points, and percentile scores improved by 13.5 points.

**Figure 2:**
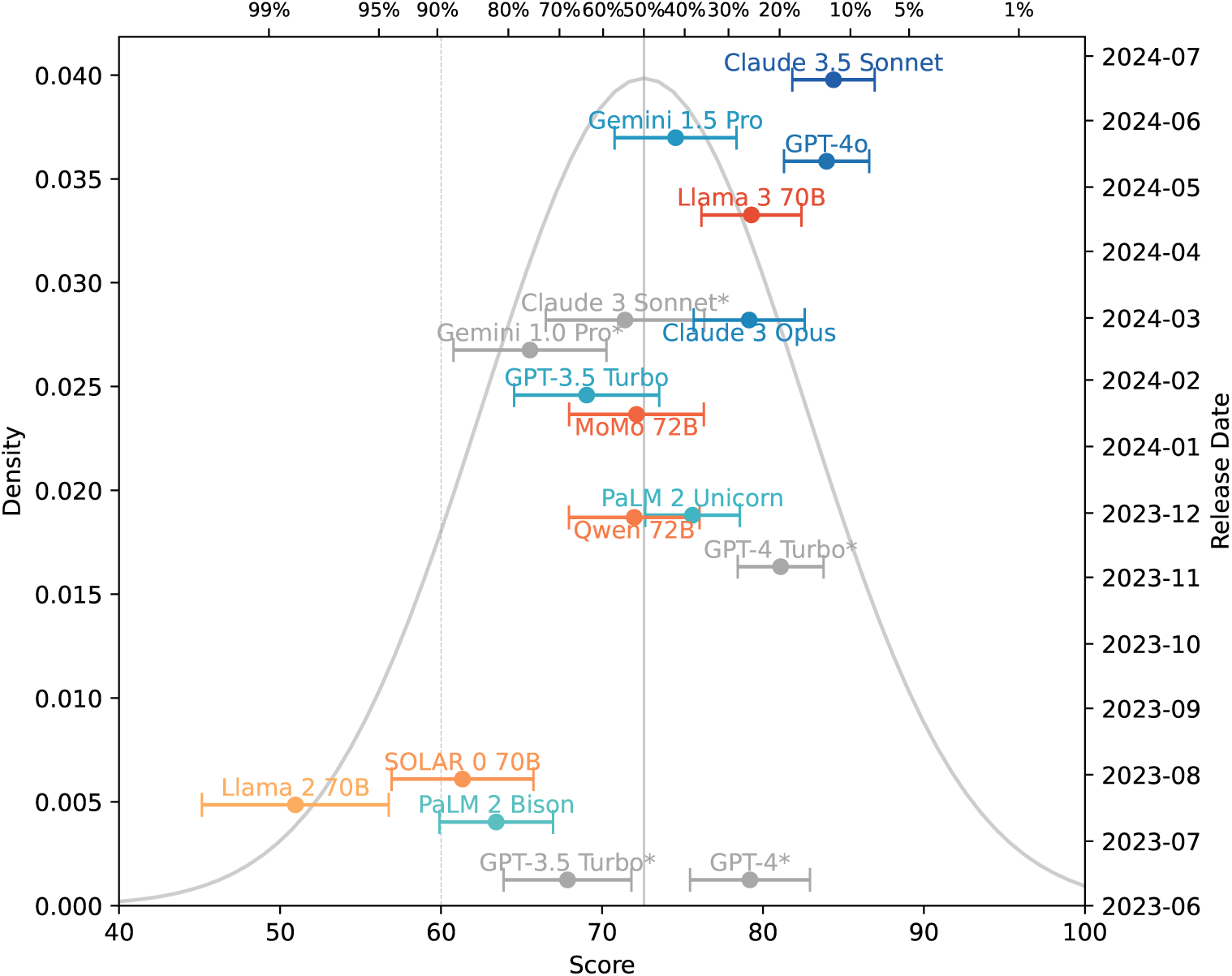
Temporal trend of LLM performance on the KPLE exam. Each point represents a specific model, showing its raw score (bottom x-axis) and percentile ranking relative to human examinees (top x-axis). Models are categorized into proprietary (blue series), open-source (red series), and legacy proprietary models (gray), with horizontal error bars indicating standard deviation across six examination years (2019-2024). The background density curve represents human score distribution, and vertical lines indicate passing thresholds (60% raw score, 50th percentile).

For the open-source models, Llama 2 70B (July 2023) initially ranked only in the top 98.5% of examinees (percentile score 1.5). In January 2024, the first open-source model MoMo 72B started to pass English-translated exams. Llama 3 70B, the best-performing open-source model as of April 2024, achieved a score placing it in the top 25.2% of examinees (percentile score 74.8). Within the approximately 12-month window from Llama 2 70B (July 2023) to Llama 3 70B (April 2024), the raw score increased by 28.3 points, and the percentile score of Llama series improved by 73.3 points.

The performance gap between open-source and proprietary models has steadily narrowed over time. Open-source models have demonstrated particularly rapid improvements. At the time of release, Llama 2 70B achieved 64.4% of the state-of-the-art performance set by GPT-4, whereas Llama 3 70B reached up to 97.9% of GPT-4 Turbo’s state-of-the-art performance at the time of its release.

### 3.3 Subject Comparison

The proprietary models generally outperformed open-source models across all subjects as illustrated in Figure 3. Within the open-source models, performance of a model is strongly correlated with its parameter size. The large models scored higher than the medium and small models (Large > Medium > Small). All model groups performed best in Biopharmacy and worst in Medical Health Legislation, while achieving similar scores in Industrial Pharmacy and Clinical & Practical Pharmacy.

**Figure 3:**
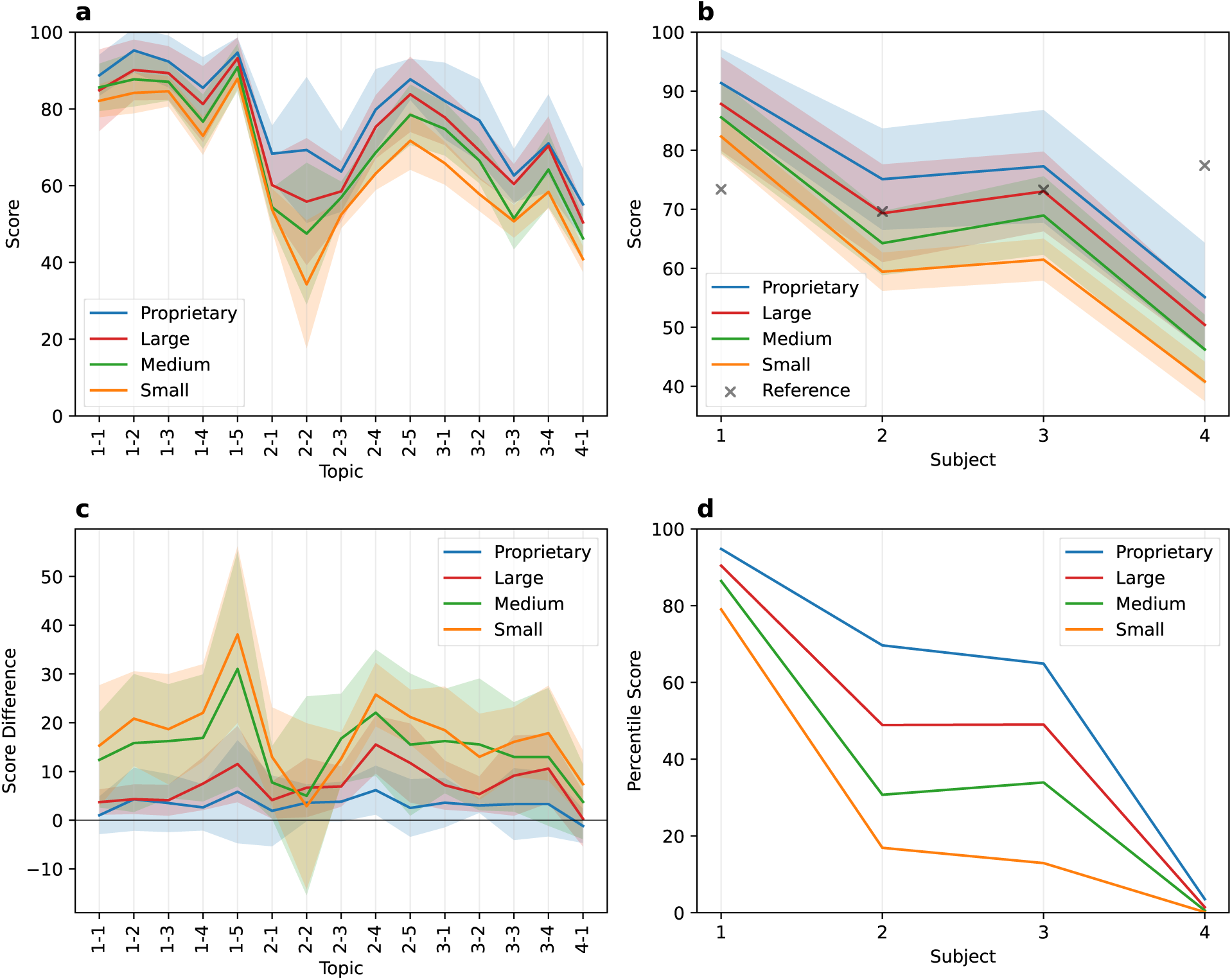
Subject-specific performance comparison among model groups. (a) Average scores per topic across different model groups: proprietary, open-source large, medium, and small models. Shaded areas indicate standard deviation within each group. (b) Average scores per subject for each model group, with human performance included as reference points (marked with “×”) for comparison. (c) Differences in scores between the English and Korean versions of exam (English–Korean) per subject for each model group. Positive values indicate higher scores on the English exam. (d) Percentile score by subject across model groups compared to human examinees across subjects, highlighting relative strengths and weaknesses.

At the topic level, models demonstrated strong performance in the following order: Pathophysiology [1–5], Microbiology & Immunology [1–2], Pharmacology [1–3], and Pharmacognosy [2–5]. Excluding Pharmaceutical & Medicinal Chemistry [2–2], where the majority of questions were edited, models struggled most with Medical Health Legislation [4], followed by Pharmaceutical Quality Science [3–3], Pharmaceutical Analysis [2–3], and Physical Pharmacy [2–1]. Additionally, differences in accuracy between the original Korean and English-translated exams were smaller among higher-scoring model groups. Notably, all model groups reached percentile scores close to 90 in Biopharmacy, whereas their percentile scores in Medical Health Legislation approached nearly zero. Even the highest-performing model, GPT-4o, achieved only a percentile score below 20 in Subject 4 (Supplementary Figure 1d), reflecting the particular challenge that nation-specific legal knowledge poses for current LLMs.

While subjects vary in the number of questions (Table 3), the total possible score is standardized across all subjects. The similar percentile scores observed between Industrial Pharmacy [2] and Clinical & Practical Pharmacy [3] suggest that differences in question counts do not systematically bias subject-specific failure rates. However, Medical Health Legislation [4] exhibited distinctly lower scores, making it the sole subject responsible for all subject-specific failures. This pattern indicates that poor performance in Subject 4 represents a fundamental limitation in current LLMs’ handling of nation-specific regulatory content rather than an artifact of examination structure.

The subject-specific development trends depicted in Supplementary Figure 1 indicate improvements in model performance across all four subjects over time. Compared to human score distributions, models exhibited excellent performance in Biopharmacy, intermediate performance in Clinical & Practical Pharmacy, and relatively poor performance in Industrial Pharmacy and Medical Health Legislation. These subject-specific performance trends among models closely resembled their overall exam performance patterns.

### 3.4 Accuracy and Difficulty Analysis

Examining the distribution of error rates at the individual item level, the low-scoring model group exhibited higher error rates compared to the high-scoring group, although the overall trends were similar (Figure 4a-b). The frequency of errors generally declined as item difficulty decreased, with edited questions making up a greater proportion of items frequently missed by multiple models. Interestingly, some edited items were answered correctly by numerous models. These questions are typically solvable without the edited content by examinees familiar with the subject matter (Supplementary Table 3).

**Figure 4:**
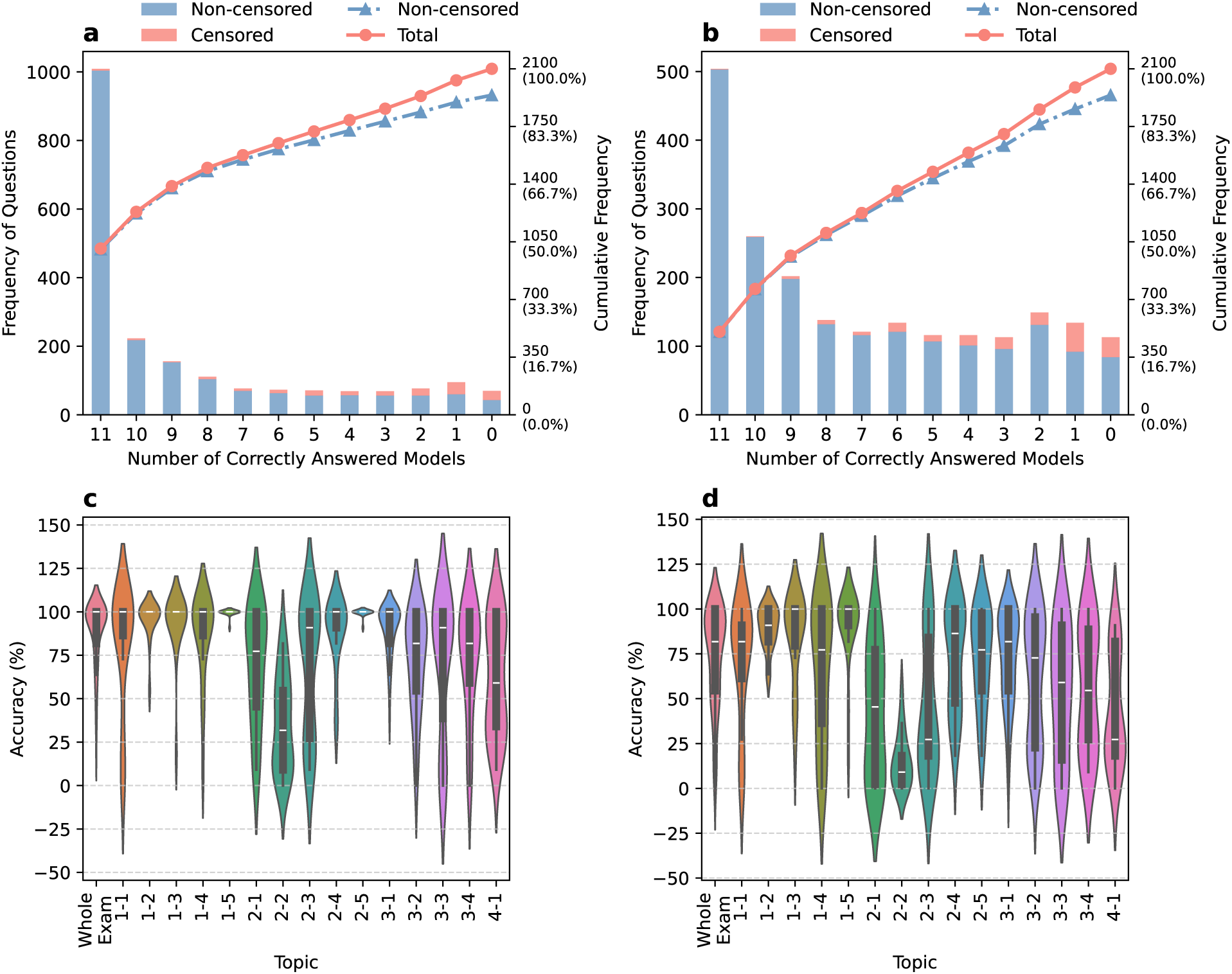
Item-level accuracy distributions of LLMs on the KPLE exam. (a–b) Frequency distribution of correct responses for unedited (blue) and edited (red) questions in (a) high-scoring and (b) low-scoring model groups. Points and bars represent the number of models answering correctly, with cumulative frequency and percentage shown on the y-axis. (c–d) Per-item accuracy distribution by topic for high-scoring (c) and low-scoring (d) groups. Violin plots show the distribution of individual question accuracy rates across topics, with each data point representing a single question’s accuracy rate within its topic. It should be noted that minor rendering artifacts may cause violin boundaries to slightly exceed the 0-100% range.

Per-item accuracy analysis across topics revealed that both high-scoring and low-scoring groups showed consistent performance patterns (Figure 4c-d). The high-scoring group achieved median accuracy rates exceeding 80% across most topics, with the narrowest distributions observed in Biopharmacy topics and the widest in topics requiring complex calculations such as Physical Pharmacy and Pharmaceutical Analysis. The low-scoring group exhibited similar topic-wise patterns but with generally lower accuracy rates and wider distributions, indicating greater variability in question-level performance. Notably, both groups demonstrated comparable accuracy distributions in the “Whole Exam” category, suggesting that the overall difficulty trends are consistent regardless of model performance level. These findings confirm that topic-specific challenges identified in subject-level analyses reflect genuine differences in question difficulty rather than artifacts of model selection or grouping methodology.

The distribution of item difficulty followed a similar trend to previous analyses (Figure 5). LLMs showed proficiency in the following subject order: Biopharmacy [1] > Industrial Pharmacy [2], Clinical & Practical Pharmacy [3] > Medical Health Legislation [4]. At the topic level, LLMs performed best in Microbiology & Immunology [1–2], Pathophysiology [1–5], Pharmacology [1–3], and Pharmacognosy [2–5]. Excluding the heavily edited Pharmaceutical & Medicinal Chemistry [2–2], models performed poorly in Medical Health Legislation [4], Pharmaceutical Analysis [2–3], Pharmaceutical Quality Science [3–3], and Physical Pharmacy [2–1]. In contrast, human examinees performed best in Medical Health Legislation [4], followed by Biopharmacy [1], Clinical & Practical Pharmacy [3], and lastly Industrial Pharmacy [2].

**Figure 5:**
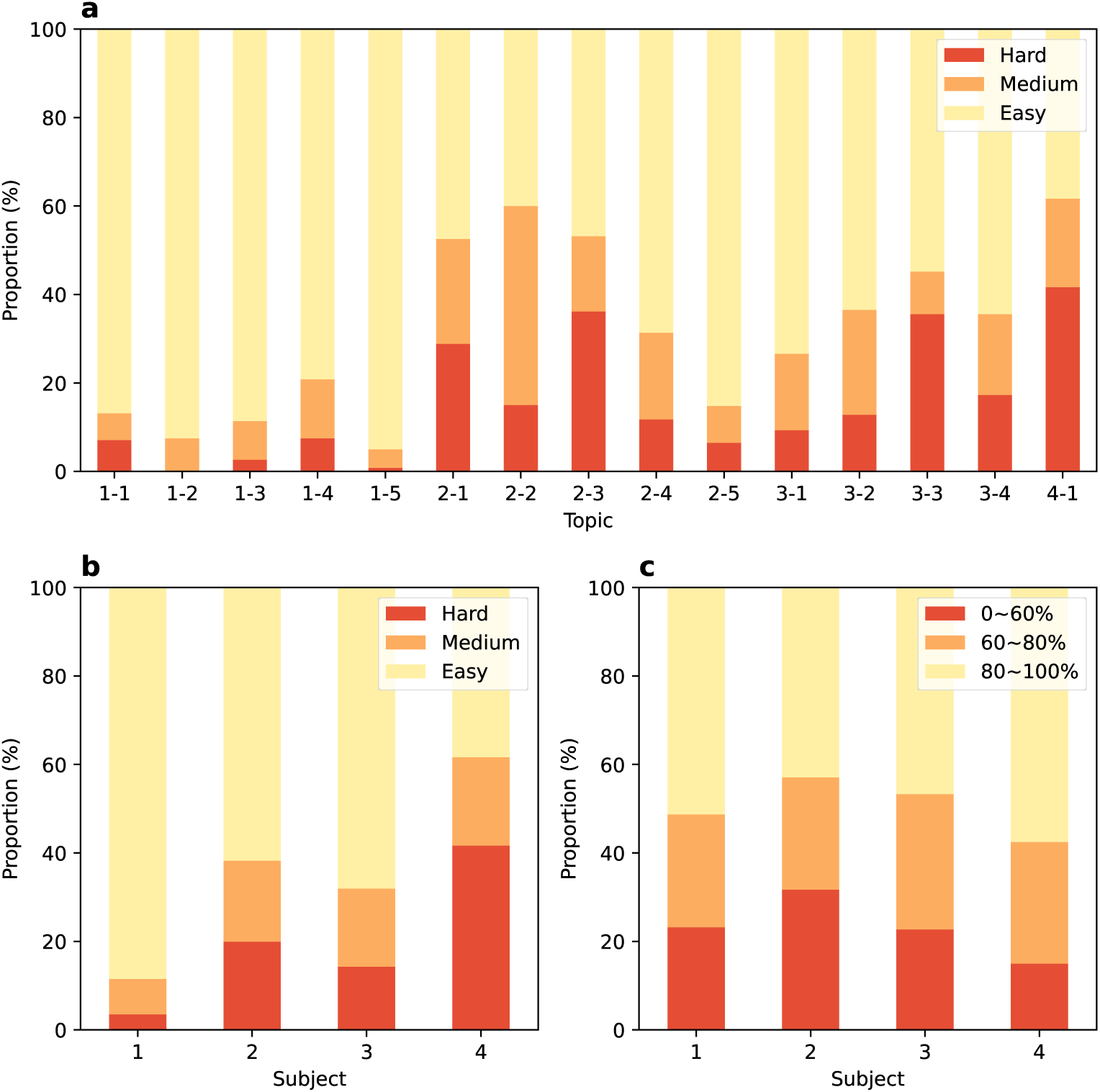
Item difficulty distribution of the high-scoring group based on model accuracy. (a) Proportion of items categorized as easy (light yellow), medium (orange), and hard (red) per individual topic. (b) Difficulty distribution by subject, following the same categorization. (c) Difficulty distribution based on actual examinee responses, categorized by KHPLEI according to the correct response rate of each item.

The difficulty distribution of the low-scoring group closely resembled that of the high-scoring group (Supplementary Figure 2). When comparing difficulty patterns between the two groups, 352 items (18.1%) were categorized as Low>High (more difficult for the low-scoring group), 1,573 items (81.1%) as LowHigh (similar difficulty), and only 15 items (0.8%) as Low<High (more difficult for the high-scoring group). This high proportion of LowHigh items indicates that both groups exhibit similar difficulty trends across the examination.

Analysis of difficulty comparison patterns across topics revealed no clear relationship with item type or specific topics, with the exception of Pharmaceutical & Medicinal Chemistry [2–2] (Figure 6). When topic 2-2 was excluded from the analysis, items from all difficulty comparison categories (Low>High, LowHigh, and Low<High) were distributed relatively evenly across different item types and topics, suggesting that the difficulty patterns between high-scoring and low-scoring groups are consistent regardless of content area or question format.

**Figure 6:**
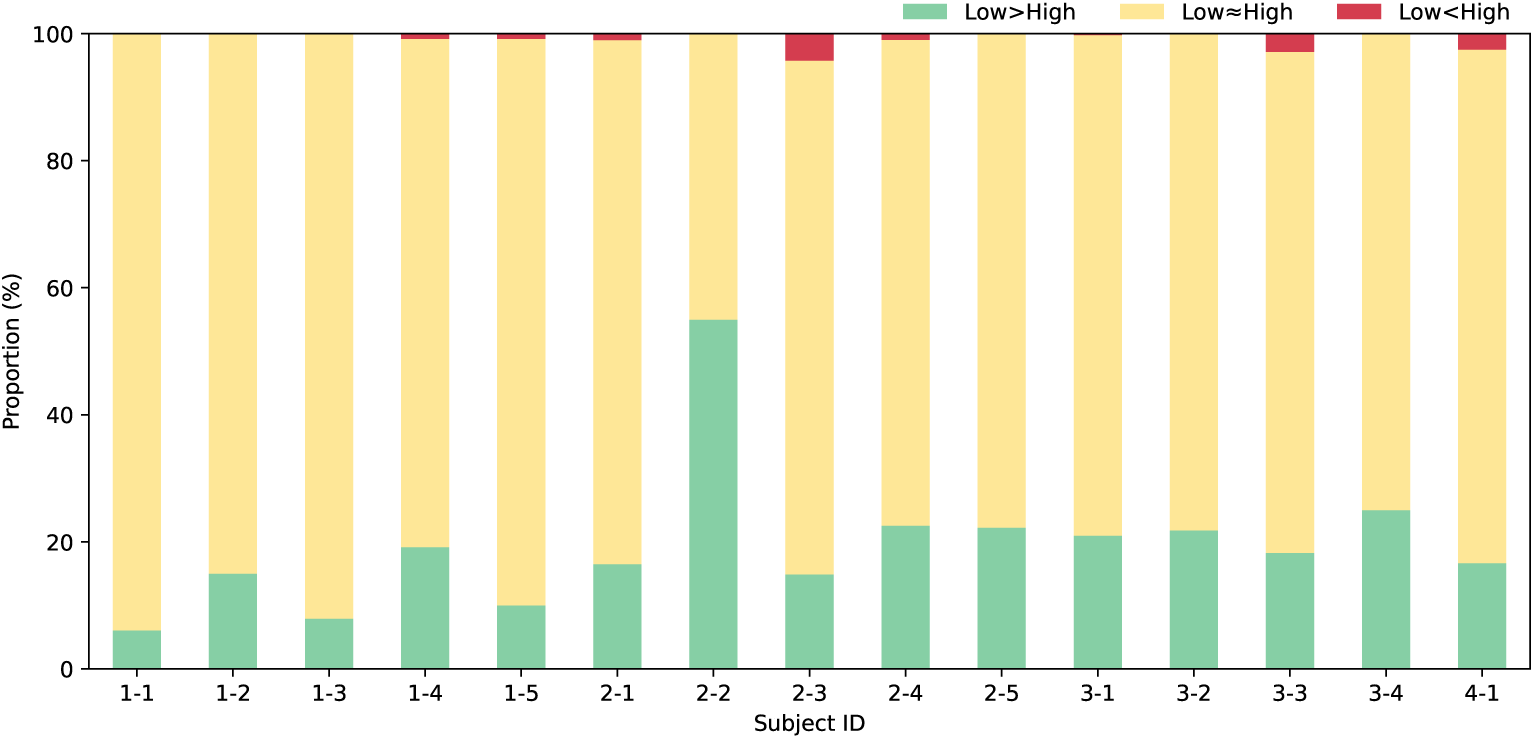
Comparison of difficulty patterns between high-scoring and low-scoring model groups. The stacked bar chart shows the proportion of items in each topic categorized by difficulty comparison: Low>High (green, items more difficult for low-scoring group), Low*≈*High (yellow, similar difficulty across groups), and Low<High (red, items more difficult for high-scoring group). Topic 2-2 (Pharmaceutical & Medicinal Chemistry) shows exceptional patterns due to the high proportion of edited questions.

## 4 Discussion

### 4.1 LLM Performance Analysis and Optimization Factors

#### 4.1.1 Parameter Size and Performance

The most evident factor influencing model performance in this study was the number of parameters. Our analysis indicated a clear trend where models with larger parameter sizes generally achieved higher accuracy. For instance, the Llama 3 70B model achieved a score of 65.3, which was 14.0 points higher—and corresponded to a percentile score increase of 51.4—compared to the smaller Llama 3 8B model from the same model family. This highlights the significant role of parameter size in enhancing model performance.

However, when comparing models of similar parameter sizes, we observed that performance improvements tend to follow a logarithmic function relative to parameter count. This suggests diminishing returns in performance gains as parameter sizes grow larger, indicating that merely increasing the number of parameters may not guarantee indefinite improvements in model accuracy. Furthermore, increasing the parameter size substantially raises the required GPU memory and computational cost. Thus, larger parameter sizes may not necessarily translate to better performance in practical applications. Therefore, selecting models with high performance-to-parameter ratios is critical for balancing accuracy and computational efficiency.

#### 4.1.2 Advancements in Model Architecture and Dataset Quality

Our results suggest that model performance improvement is not solely driven by increases in parameter size. Two additional critical factors, advancements in model architecture and improvements in dataset quality, contribute significantly to performance enhancement. While it is challenging to isolate and analyze these two effects independently, their interplay clearly played a critical role in overall model performance advancements.

For example, Llama 3 70B, the successor of Llama 2 70B, improved substantially, showing an increase of 28.3 points in raw score and 73.3 points in percentile score. Additionally, recent trends have demonstrated that newer models with fewer parameters can outperform older models with larger parameter sizes. Gemma 2 27B and Phi 3 Medium exemplify this trend, achieving high efficiency with smaller parameter sizes. The structural modifications of existing model have also yielded significant improvements, as demonstrated by SOLAR 10.7B, which was developed through depth up-scaling of the Mistral 7B architecture [35].

With the incorporation of diverse multilingual datasets and improvements in dataset quality, recent LLMs demonstrate significantly reduced performance gaps across languages. Our study also shows that the performance gap between English and Korean has decreased. This trend is consistent with findings from recent studies indicating that newer models with enhanced training data diversity exhibit improved robustness and fairness across a broader range of languages, including low-resource ones [64].

Future advancements in both dataset accumulation and model architecture will lead to improvement of LLM performance. Notably, next-generation architectures such as Mamba are actively being investigated to improve the conventional transformer architecture [65].

#### 4.1.3 Cross-Linguistic Performance and Translation Effects

Our findings of reduced performance gaps between Korean and English exam versions align with improved multilingual capabilities observed across non-English pharmacist examinations. Kunitsu et al.[13] demonstrated GPT-4’s effective performance on Japanese examination questions without translation, achieving 78% accuracy on answerable items. Wang, Shen, and Chen [15] quantified substantial language-dependent performance differences in Taiwan’s examination, where GPT-3.5 scored 54% in Chinese versus 57% in English for basic science questions, with gaps widening to 30-37% for clinical subjects. Zong et al. [16] observed similar patterns in Chinese medical examinations, noting that while language barriers affected earlier models substantially, GPT-4 demonstrated improved cross-linguistic consistency. Jin and Kim [17] reported GPT-4 achieving 86% accuracy on Korean examination, confirming effective multilingual performance.

These cross-study comparisons reveal convergent findings: (1) consistent performance gaps favoring English versions in earlier models (GPT-3.5), (2) progressive reduction in language-dependent performance differences with architectural advances (GPT-4 and beyond), and (3) persistent challenges in clinical and culturally-specific content despite improved multilingual training. Our observation that certain models (e.g., Yi 1.5 34B, Phi 3 Medium) passed all English exams but failed Korean versions reflects this ongoing challenge, though the gap narrows with each model generation.

Direct comparison with Jin and Kim [17], which evaluated GPT models on identical Korean examination materials 12 months prior, reveals remarkable progress. Current open-source models now achieve performance levels previously exclusive to proprietary models, demonstrating both rapid architectural advancement and expanding multilingual training data diversity [64].

#### 4.1.4 Influence of Topics and Question Types

LLMs performed best in Biopharmacy and worst in Medical Health Legislation, with intermediate results in Industrial Pharmacy and Clinical & Practical Pharmacy. They excelled in Microbiology & Immunology [1–2], Pharmacology [1–3], Pathophysiology [1–5], and Pharmacognosy [2–5], but struggled with Physical Pharmacy [2–1], Pharmaceutical Analysis [2–3], Pharmaceutical Quality Science [3–3], and Medical Health Legislation [4].

Strong performance is correlated with subjects emphasizing factual recall, aligning with findings by Xie et al. [66], which demonstrated that LLMs tend to excel when solving problems by retrieving memorized information. In contrast, topics with low performance—Physical Pharmacy and Pharmaceutical Analysis—require complex calculations or reasoning to solve problems, which LLMs find more challenging.

It is noticeable that Pharmaceutical Quality Science and Medical Health Legislation were challenging despite their ostensibly knowledge-based nature, possibly due to their nation-specific nature of topics. Pharmaceutical Quality Science requires familiarity with Korean Pharmacopeia (KP) and Korea Good Manufacturing Practice (KGMP), while Medical Health Legislation demands an understanding of Korean laws and regulations. This difficulty shows that the current LLMs may suffer from insufficient training on nation-specific standards and regulations. These results suggest that although LLMs are expected to be trained on broad datasets, they may struggle with topics requiring specialized and country-specific knowledge, especially laws and regulations, which are critical in medical and pharmaceutical practices. Thus, caution is advised when employing LLMs in these domains. Overall, LLMs excel in areas requiring factual recall but falter in domains demanding complex reasoning or highly specific, region-dependent knowledge.

#### 4.1.5 Fine-tuning

In this study, we observed clear performance improvements in fine-tuned models. For example, SOLAR 0 70B, a fine-tuned version of Llama 2 70B, exhibited significant gains in both Korean and English scores. On the other hand, MoMo 72B, derived from fine-tuning Qwen 72B, improved its performance primarily on Korean questions, thus reducing the gap between English and Korean scores; however, its English scores remained largely unchanged. This result illustrates that fine-tuning selectively enhances performance depending on the characteristics of the datasets used.

Consequently, the choice of an appropriate fine-tuning dataset emerges as a critical factor in determining the success of fine-tuning. Incorporating linguistically diverse data or datasets specifically tailored to address gaps in the model’s knowledge can effectively drive targeted performance improvements. Specifically, given that our study identified poor LLM performance in topics such as Pharmaceutical Quality Science [3–3] and Medical Health Legislation [4], we anticipate significant reductions in performance gaps if models were fine-tuned using datasets containing detailed, country-specific content from these areas, such as the Korean Pharmacopeia (KP), Korea Good Manufacturing Practice (KGMP), and relevant laws and regulations.

#### 4.1.6 Comparison with Human Performance

LLMs demonstrated a clear strength in Biopharmacy, whereas their weakest performance was observed in Medical Health Legislation. In contrast, human examinees performed best in Medical Health Legislation while generally finding Industrial Pharmacy to be the most challenging. These results further confirm that LLMs have limited accuracy in answering nation-specific legal questions, which are featured in Medical Health Legislation. However, LLMs out-perform human examinees on memorization-oriented questions, consistent with LLMs’ primary capabilities of pattern recognition and information processing derived from large-scale textual data.

The observed difficulty patterns—LLMs excelling in memorization-based topics but struggling with nation-specific regulations—are consistent with findings from pharmacist examinations in other countries [13, 17]. Notably, recent models can now pass without excluding visual content questions, unlike earlier evaluations [13].

Our comparison between high-scoring and low-scoring model groups revealed remarkably similar difficulty trends, with 99.2% of items showing consistent difficulty patterns across both groups. This consistency suggests that item difficulty is an inherent property of the questions themselves rather than model-dependent factors. The exceptions—items where difficulty patterns diverged between groups—were distributed evenly across topics and item types (excluding the heavily edited Pharmaceutical & Medicinal Chemistry [2–2]), indicating no systematic bias in how different model performance levels interact with specific content domains. This finding supports the reliability of our difficulty categorization and suggests that the fundamental challenges posed by KPLE questions remain consistent regardless of model sophistication.

Humans, however, not only acquire the necessary knowledge to answer KPLE questions but also learn how to apply and utilize that knowledge effectively during exam preparation. The models evaluated in this study displayed impressive performance despite not undergoing specialized fine-tuning tailored to the KPLE. Thus, it can be expected that LLM performance could be further improved by fine-tuning the models on datasets that incorporate not only scientific knowledge but also data reflecting the problem-solving strategies and processes employed by human examinees.

#### 4.1.7 Proprietary vs. Open-Source Models

Proprietary models, such as GPT-4o and Claude 3.5 Sonnet, demonstrated outstanding performance on the translated exams, successfully passing all tests and remaining ahead of the latest open-source models. However, open-source models are rapidly advancing. For example, Llama 3 70B achieved 94% of the performance level of proprietary models. This trend indicates a narrowing performance gap between proprietary and open-source models, suggesting that open-source models may already be sufficiently competitive for specific practical applications.

Proprietary models often utilize cloud-based architectures, transferring user data to external servers for processing. This approach carries inherent risks regarding sensitive data, including potential breaches of confidentiality, a significant concern particularly in medical contexts. Conversely, open-source models can operate on local servers, creating an environment in which data remains secure and under user control. In other words, open-source models have a distinct advantage in terms of security and data governance.

Proprietary models typically function within closed ecosystems, limiting user customization and flexibility. In contrast, open-source models offer complete accessibility and flexibility, empowering users to modify or optimize models according to their specific needs and tasks, thereby providing greater flexibility and adaptability.

### 4.2 LLM & Pharmacist

#### 4.2.1 Implications of LLMs Passing the KPLE

The ability of LLMs to pass the Korean Pharmacist Licensing Examination (KPLE) does not imply that these models can replace pharmacists in practice. Pharmacists possess specialized expertise developed not only through acquiring academic knowledge but also through practical training and clinical experience. The KPLE assesses essential knowledge required for pharmaceutical practice, rather than directly evaluating practical pharmaceutical care. Moreover, the pharmacist’s role transcends mere knowledge delivery, involving critical human interactions such as patient communication, clinical judgment, and ethical decision-making. Currently, LLMs lack the emotional intelligence and interpersonal skills required for these human interactions, and therefore cannot fully substitute the professional expertise and responsibilities of pharmacists. Thus, LLMs passing the KPLE do not indicate that pharmacists are at risk of being replaced by AI.

Additionally, the accuracy and reliability of information provided by LLMs require careful verification. Given the critical nature of pharmaceutical practice, any inaccuracies in AI-generated information could lead to serious consequences. Therefore, LLMs should be utilized as complementary tools to pharmacists, with the ultimate responsibility and clinical judgment remaining firmly under human oversight.

It is also essential to initiate discussions about incorporating assessments of pharmacist competencies that AI cannot replicate into licensing exams. For example, evaluating skills such as patient communication, clinical decision-making, and ethical judgment—areas in which current LLMs clearly fall short—will become increasingly important in distinguishing human expertise from AI.

#### 4.2.2 Utilizing LLMs to Enhance Pharmacists’ Professional Expertise

Although current LLMs exhibit limitations in high-level tasks requiring complex reasoning and clinical judgment, they already surpass human performance in memorization-based tasks and text-based information processing. Considering these strengths, delegating repetitive and routine pharmaceutical tasks to LLMs could allow pharmacists to concentrate more on higher-level, specialized activities, potentially improving their professional expertise and overall work efficiency.

LLMs open possibilities for AI-driven tools to support pharmaceutical education, either as learning aids or content development tools. For instance, students can utilize LLMs as educational copilots to facilitate understanding of complex pharmaceutical concepts. Additionally, simulated examinations powered by LLMs can help students regularly assess their knowledge, enhancing learning efficiency. Currently, these systems are particularly effective for memorization-oriented content, but future improvements in LLM capabilities may enable the generation of complex clinical scenarios, further contributing to the enhancement of students’ clinical reasoning skills.

LLMs can also serve as powerful tools for data analysis, information retrieval, and document preparation. For example, LLMs could support pharmacists during drug discovery by facilitating literature reviews and data analyses or could rapidly supply relevant information to hospital pharmacists when reviewing patient-specific medication regimens. Moreover, pharmacists could leverage LLMs for analyzing complex drug interactions and pharmacological data, thereby enhancing efficiency and enabling more accurate clinical decision-making.

When integrating LLMs into pharmacy practice, factors such as model performance, security, costs, and scalability should be comprehensively considered. Proprietary models provide high performance and ease of use but raise concerns regarding data security and substantial long-term costs. Conversely, open-source models offer greater security and flexibility, though initial investments and ongoing management costs may be significant. Therefore, it is crucial to select appropriate models based on the specific use cases and environmental requirements of pharmacy practice. Particularly when choosing open-source models, careful evaluation of performance-to-resource efficiency (performance-parameter ratio) is essential to determine the most suitable model for deployment.

### 4.3 Limitations

#### 4.3.1 Limitations of the Study’s Prompting Strategy

The use of zero-shot prompting in this study likely limited the models’ ability to demonstrate their full potential. This approach is akin to asking a person to solve a complex problem without the opportunity to break it down into smaller steps or recall relevant information explicitly. Specifically, the zero-shot prompting strategy used here, which only requested the correct answer number, might not have fully utilized the reasoning and problem-solving capabilities of the models. Existing research indicates that Chain of Thought (CoT) prompting, which encourages models to “think step by step,” can significantly enhance their performance by guiding them through a structured reasoning process [67, 68]. As such, it is plausible that employing these more advanced prompting strategies could lead to substantially better performance than reported in this study.

#### 4.3.2 Comparison with Human Examinees

KHPLEI does not publicly release item-level data, limiting direct model-human comparisons. Specifically, individual question performance metrics (e.g., correct response rates per item) and question type classifications remain confidential. This data gap prevents nuanced analysis of which specific question types pose greater challenges for models versus humans.

While KHPLEI categorizes questions into three cognitive types (Recall/Interpretation/Problem-solving), item-level classifications distinguishing knowledge-based (recall/comprehension) from skills-based (calculation/interpretation) questions were not provided. Rigorous manual classification of all 2,100 questions was not feasible due to inherent ambiguities and unavoidable subjectivity. However, our subject-level performance analysis indirectly addresses these cognitive demands by demonstrating that LLMs excel in memorization-intensive subjects while struggling with calculation-intensive domains, providing meaningful insights into model capabilities across different cognitive requirements. Future collaborations with educational institutions could help obtain more granular data, enabling more precise adjustments and improvements in model training.

#### 4.3.3 Impact of Edited Questions

The exclusion of visual content from 7.6% of questions (160/2,100), particularly affecting Industrial Pharmacy subjects such as Pharmaceutical & Medicinal Chemistry (81.5% exclusion rate), limits our evaluation to text-based problem-solving capabilities. While multimodal models with vision capabilities have been available since GPT-4V [24], this study focused on standardized text-only evaluation for consistent comparison across diverse model architectures. However, this approach may underestimate the true capabilities of current LLMs in pharmaceutical contexts. Future studies should incorporate multimodal evaluation using original visual content to assess models’ ability to interpret chemical structures, reaction mechanisms, and analytical data—essential skills for pharmaceutical practice.

#### 4.3.4 Context Within the Research Landscape

Our comprehensive benchmarking study contributes to the growing body of evidence on LLM performance in pharmaceutical licensing contexts while addressing previous gaps in systematic comparison across model architectures. Prior studies consistently demonstrated GPT-4’s capability to pass pharmacist examinations: 78% accuracy in Japan [13, 14], 68% in Taiwan (English version) [15], 86% in Korea [17], while GPT-3.5 consistently failed to achieve passing scores across all evaluated contexts. These studies collectively identified common performance patterns—superior accuracy in pharmacology and basic sciences, persistent weaknesses in calculation-intensive topics and nation-specific regulations—that align with our subject-level findings.

Our study advances this research landscape through three distinctive contributions. First, systematic evaluation of 27 models spanning proprietary and open-source architectures reveals that current open-source models (e.g., Qwen2 72B, Llama 3 70B, Gemma 2 27B) now achieve passing performance previously limited to proprietary systems, demonstrating rapid democratization of pharmaceutical knowledge capabilities. Second, temporal analysis across 12-month development period quantifies unprecedented improvement rates, particularly among open-source models, providing empirical evidence for accelerating capability advancement. Third, detailed subject-level analysis across 2,100 questions with consistent difficulty patterns between high- and low-performing model groups establishes that performance limitations reflect inherent question characteristics rather than model-specific factors, validating the robustness of our difficulty categorization. As with prior examination-specific studies, our findings reflect KPLE’s particular structure and content, requiring appropriate caution in generalization to other contexts.

## 5 Conclusion

This comprehensive benchmarking study evaluated 27 LLMs across multiple dimensions—including proprietary and open-source architectures, parameter sizes ranging from 4B to 104B, multiple model generations, and both Korean and English language versions—providing systematic insights into LLM capabilities on the Korean Pharmacist Licensing Examination. Our findings demonstrate that current state-of-the-art models consistently achieve passing scores, with performance driven by multiple interacting factors including parameter scaling, architectural innovations, multilingual training data quality, and fine-tuning strategies.

Temporal analysis reveals rapid performance improvement across all model categories, with particularly notable advances in open-source models narrowing the performance gap substantially over the 12-month study period. Size-based comparison shows logarithmic returns on parameter scaling, while architectural innovations enable recent smaller models to surpass older larger predecessors. Cross-linguistic evaluation demonstrates improving multilingual capabilities in newer generations, though performance disparities persist in certain model families.

Subject-level analysis identifies systematic performance patterns: models excel in memorization-intensive domains (Biopharmacy, Pharmacology) while showing consistent limitations in areas requiring complex calculations (Physical Pharmacy, Pharmaceutical Analysis) and nation-specific knowledge (Medical Health Legislation, Pharmaceutical Quality Science). These findings indicate clear directions for targeted improvement through domain-specific fine-tuning, specialized training datasets incorporating regional standards and regulations, and enhanced reasoning capabilities.

These results emphasize the considerable potential of LLMs as supportive tools for pharmacy education and professional practice. Future research should prioritize developing specialized fine-tuning strategies to address identified performance gaps, exploring advanced prompting techniques beyond zero-shot approaches, and investigating the ethical, regulatory, and educational implications of LLM integration into pharmaceutical practice. Both proprietary and open-source models offer distinct advantages—high performance and ease of use versus security, flexibility, and customizability—suggesting that model selection should be guided by specific use cases and environmental requirements.

Ultimately, fostering a collaborative human-AI model will leverage the complementary strengths of pharmacists’ clinical expertise and LLMs’ information processing capabilities, enhancing professional efficiency while maintaining the critical human oversight necessary for safe pharmaceutical practice.

## 6 List of abbreviations

AI: Artificial Intelligence
LLMs: Large Language Models
KPLE: Korea Health Personnel Licensing Examination
KHPLEI: Korea Health Personnel Licensing Examination Institute

## 7 Supplementary information

- Title: Supplementary Materials

- File name: supplementary.pdf
- Contents: Supplementary Tables 1–3, Figure 1-2

## Declarations

- This work was supported by Seoul National University (370C-20220109 and AI-Bio Research Grant 0413-20230053), the National Research Foundation of Korea (Grant nos. RS-2023-00256320, 2022M3E5F3081268, and 2022R1C1C1005080), and the Institute of Information & Communications Technology Planning & Evaluation (IITP) grant funded by the Korea government (MSIT) (RS-2023-00220628, Artificial intelligence for prediction of structure-based protein interaction reflecting physicochemical principles). This research was also supported by the Bio & Medical Technology Development Program of the National Research Foundation (NRF) funded by the Korean government (MSIT) (No. RS-2024-00352229). This work was also supported by the Korea Drug Development Fund funded by the Ministry of Science and ICT, Ministry of Trade, Industry, and Energy, and Ministry of Health and Welfare (RS-2023-00217308). This work was supported by Korea Environment Industry & Technology Institute(KEITI) through “Advanced Technology Development Project for Predicting and Preventing Chemical Accidents” Program, funded by Korea Ministry of Environment(MOE)(RS-2023-00219144).
- Competing interests: Not applicable.
- Consent for publication: Not applicable.
- Data availability: All datasets generated and analyzed in this study (including model-generated answers, scoring results, and performance analyses) are publicly available in the Harvard Dataverse: https://doi.org/10.7910/DVN/YCXPT5. However, the original KPLE datasets and processed exam texts are not included in this repository due to copyright restrictions. Original exam texts were obtained from the Korea Health Personnel Licensing Examination Institute (KHPLEI) and are publicly available at: https://www.kuksiwon.or.kr/CollectOfQuestions/brd/m_116/list.do.
- Materials availability: Not applicable.
- Code availability: All code used in this study is publicly available in the Github repository: https://doi.org/10.5281/zenodo.15209032.
- Author contribution: D.H.J.: Conceptualization, Investigation, Methodology, Data curation, Formal analysis, Software, Visualization, Writing – Original draft, Writing – Review & Editing. J.L.: Conceptualization, Supervision, Resources, Funding acquisition, Writing – Review & Editing. All authors read and approved the final manuscript.

