## Supplementary Table 1 for "Proprietary and Open-Source Large Language Models on the Korean Pharmacist Licensing Examination: A Comparative Benchmarking Study"

### Supplementary Materials

Table 1: Comparison of response formats from `Llama-2-70b-hf` and `Llama-2-70b-chat-hf` to a sample KPLE question. Only the chat-optimized model provided the correct answer in the required format.

| Model | Llama-2-70b-hf | Llama-2-70b-chat-hf |
|---|---|---|
| Prompt: | You are taking the Pharmacist Licensing Examination. Read the question carefully and use your judgment to select one of the options beginning with '①, ②, ③, ④, ⑤' as an answer. Submit only one symbol from '①, ②, ③, ④, ⑤' without any explanation, as this is an OMR exam.<br><br>58. Which drug is used for metastatic colorectal cancer and metastatic non-small cell lung cancer by binding to vascular endothelial growth factor (VEGF), thereby interfering with VEGF binding to its receptor?<br><br>① Imatinib<br>② Erlotinib<br>③ Cetuximab<br>④ Bevacizumab<br>⑤ Panitumumab | <s> [INST] «SYS»<br>You are taking the Pharmacist Licensing Examination. Read the question carefully and use your judgment to select one of the options beginning with '①, ②, ③, ④, ⑤' as an answer. Submit only one symbol from '①, ②, ③, ④, ⑤' without any explanation, as this is an OMR exam.<br>«/SYS»<br><br>58. Which drug is used for metastatic colorectal cancer and metastatic non-small cell lung cancer by binding to vascular endothelial growth factor (VEGF), thereby interfering with VEGF binding to its receptor?<br><br>① Imatinib<br>② Erlotinib<br>③ Cetuximab<br>④ Bevacizumab<br>⑤ Panitumumab [/INST] |
| Response: | 59. Which of the following is a drug that is used to treat chronic myeloid leukemia by inhibiting the tyrosine kinase activity of BCR-ABL?<br><br>① Imatinib<br>② Erlotinib<br>③ Cetuximab<br>④ Bevacizumab<br>⑤ Panitumumab<br><br>60. Which of the following is a drug that is used to treat chronic myeloid leukemia by inhibiting the tyrosine kinase activity of BCR-ABL?<br><br>① Imatinib<br>② Erlotinib<br>③ Cetuximab<br>④ Bevacizumab<br>⑤ Panitumumab<br><br>61. ... | Sure, I can help you with that!<br><br>My answer is ④ Bevacizumab.</s> |
