## Supplementary Table 2 for "Proprietary and Open-Source Large Language Models on the Korean Pharmacist Licensing Examination: A Comparative Benchmarking Study"

Table 2: Results of preliminary tests comparing translation quality on the KPLE using different language models

(a) First-round results comparing DeepL and GPT-4-translated exams (October 2023)

| **Model** | **DeepL (10/5/2023)** | **gpt-4-0613** |
|---|---|---|
| gpt-4-0613 | 74.7 | 77.7 |
| gpt-3.5-turbo-0613 | 66.3 | 66.1 |
| SOLAR-0-70b-16bit | 56.1 | 59.5 |
| chat-bison@001 | 49.5 | 52.4 |
| Llama-2-70b-chat-hf | 46.0 | 46.8 |

(b) Second-round results comparing GPT-4 Turbo and Claude 3 Opus translations (May 2024)

| **Model** | **gpt-4-turbo-2024-04-09** | **claude-3-opus@20240229** |
|---|---|---|
| gpt-4-turbo-2024-04-09 | 80.7 | 81.5 |
| gpt-3.5-turbo-0125 | 68.3 | 69.7 |
| claude-3-opus@20240229 | 80.7 | 81.6 |
| Meta-Llama-3-8B-Instruct | 60.3 | 62.8 |
| SOLAR-10.7B-Instruct-v1.0 | 60.8 | 61.8 |
