## Supplementary Table 3 for "Proprietary and Open-Source Large Language Models on the Korean Pharmacist Licensing Examination: A Comparative Benchmarking Study"

Table 3: Selected Examples of KPLE Questions

| Question | Translated Text |
|---|---|
| **2019-1-02** [1-1] Easy (11/11) | Question: When skeletal muscle contracts, which component does the Ca2+ released from the sarcoplasmic reticulum bind to?<br><br>① actin<br>② troponin C<br>③ troponin I<br>④ troponin T<br>⑤ tropomyosin |
| **2020-1-29** [1-2] Easy (10/11) | Question: Which serum protein stabilizes the C3 convertase in the complement activation pathway initiated by cell wall components of pathogens?<br><br>① C1q<br>② C4b<br>③ immunoglobulin M<br>④ mannose-binding lectin<br>⑤ properdin |
| **2021-1-51** [1-3] Easy (9/11) | Question: Which antihypertensive drug has a weak side effect of dry cough and angioedema due to its mechanism of action of blocking the Angiotensin II type 1 (AT1) receptor?<br><br>① Ramipril<br>② Labetalol<br>③ Amlodipine<br>④ Telmisartan<br>⑤ Sodium nitroprusside |

| Question | Translated Text |
| --- | --- |
| **2023-1-80**<br>[1-4]<br>Easy<br>(11/11) | [Case 1]<br>• An epidemiological study was conducted to determine the causal relationship between smoking during pregnancy and low birth weight.<br>• 500 low birth weight infants and 500 normal weight infants born in a hospital from 2018 to 2020 were selected, and the mothers' smoking history during pregnancy was investigated.<br><br>[Case 2]<br>• An epidemiological study is being conducted to determine the causal relationship between smoking and lung cancer.<br>• Among visitors aged 40 and over from 2018 to 2020, 1,000 individuals who were not affected by lung cancer were divided into smokers and non-smokers.<br>• Follow-up studies on the incidence of lung cancer in both groups will be conducted for about 30 years until 2050.<br><br>Question: Compared to Case 2, what is a general advantage that the type of research method used in Case 1 might have?<br><br>① It has high internal validity.<br>② It is easy to calculate disease incidence rates and risk factors.<br>③ The research period is shorter and the cost is lower.<br>④ It is possible to observe the occurrence of various diseases according to exposure to risk factors.<br>⑤ The entire process from exposure to risk factors to disease occurrence can be observed. |
| **2024-1-98**<br>[1-5]<br>Easy<br>(11/11) | Question: Which ophthalmic condition can be treated with vascular endothelial growth factor (VEGF) inhibitors?<br><br>① Corneal ulcer<br>② Retinal infection<br>③ Exophthalmos<br>④ Macular edema<br>⑤ Retinal arteriosclerosis |

| Question | Translated Text |
| --- | --- |
| **2024-2-10**<br>[2-1]<br>Hard<br>(1/11) | Question: What is the molar concentration (mol/L) of a cocaine hydrochloride solution with a freezing point depression of 0.52°C? (Note: The L_iso value of cocaine hydrochloride is 3.4, and the difference between molar concentration and molal concentration is ignored.)<br><br>① 1/100<br>② 1/20<br>③ 11/168<br>④ 13/85<br>⑤ 34/99 |
| **2020-2-32**<br>[2-2]<br>Easy<br>(11/11) | Question: Which drug contains a pteridine ring?<br><br>① acyclovir<br>② tamoxifen<br>③ methotrexate<br>④ ketoconazole<br>⑤ cyclophosphamide |

| Question | Translated Text |
|---|---|
| **2020-2-53**<br>[2-3]<br>Hard<br>(3/11) | Aspirin tablets were stored for a long period under high temperature and humidity conditions, resulting in an irritating odor. To verify the purity of this drug, it was analyzed according to liquid chromatography method, and two peaks were detected. The retention times (tR) of each peak were 5.0 min and 10.0 min, and the retention time of the mobile phase (t0) was 2.5 min.<br><br>[Analysis Conditions]<br>Detector: UV spectrophotometer (measurement wavelength UV 280 nm)<br>Column: Stainless steel column packed with octadecylsilica gel for liquid chromatography, with an inner diameter of about 4 mm and a length of 30 cm<br>Mobile phase: 0.085% phosphoric acid : methanol (60 : 40), (pH 3.0)<br>Flow rate: 1.0 mL/min<br><br>Question: When the ’peak width at baseline (Wb)’ of the two separated peaks is 0.45 min and 0.55 min respectively, what is the resolution (R) between the two peaks?<br><br>① 3<br>② 4<br>③ 5<br>④ 10<br>⑤ 15 |
| **2021-2-63**<br>[2-4]<br>Easy<br>(11/11) | Question: Which of the following preservatives used in the manufacture of eye drops has improved preservative effects due to increased permeability against Pseudomonas aeruginosa and other gram-negative bacteria when sodium edetate is added?<br><br>① benzalkonium chloride<br>② butylated hydroxyanisole<br>③ propyl gallate<br>④ sodium bisulfite<br>⑤ stearyl alcohol |

| Question | Translated Text |
|---|---|
| **2022-2-75**<br>[2-5]<br>Easy<br>(11/11) | Question: Which alkaloid is commonly contained in the following medicinal herbs?<br><br>• Belladonnae Radix<br>• Daturae Folium<br>• Scopoliae Rhizoma<br><br>① atropine<br>② berberine<br>③ higenamine<br>④ quinidine<br>⑤ reserpine |
| **2024-3-28**<br>[3-1]<br>Medium<br>(5/11) | Question: A 57-year-old woman was diagnosed with hypernatremia due to nephrogenic diabetes insipidus. The lithium carbonate she had been taking was discontinued, and 5% glucose solution was administered intravenously. What is the appropriate additional medication?<br><br>[Medical history] Bipolar disorder<br>[Clinical tests]<br>Na 152 mEq/L, K 4.2 mEq/L, BUN 15 mg/dL, SCr 1.1 mg/dL<br>Plasma osmolality 312 mOsm/kg (reference range 275-300 mOsm/kg)<br>Urine osmolality 98 mOsm/kg (reference range 250-900 mOsm/kg)<br>Urine output 4,500 mL/24 hours (reference range 800-2,000 mL/24 hours)<br>Serum lithium concentration 1.4 mEq/L (reference range 0.6-1.2 mEq/L)<br>[Current medication] Lithium carbonate 600 mg twice daily<br><br>① Tolvaptan<br>② Lisinopril<br>③ Furosemide<br>④ Amiloride<br>⑤ Calcium polystyrene sulfonate |

<table>
<tr><th>Question</th><th>Translated Text</th></tr>
<tr><td>2019-4-04<br>[3-2]<br>Medium<br>(5/11)</td><td>A 54-year-old male recently started taking the following prescription medications for the treatment of essential hypertension and dyslipidemia, and for the prevention of coronary artery disease.<br><br>| Prescription medication name | Dose per administration | Frequency per day |<br>|---|---|---|<br>| Simvastatin tablet 20 mg | 1 | 1 |<br>| Aspirin tablet 100 mg | 1 | 1 |<br>| Amlodipine tablet 5 mg | 1 | 1 |<br>| Carvedilol tablet 12.5 mg | 1 | 1 |<br>| Hydrochlorothiazide tablet 25 mg | 1 | 1 |<br><br>Question: Which medication should be taken in the morning to prevent side effects?<br><br>① Aspirin<br>② Amlodipine<br>③ Simvastatin<br>④ Carvedilol<br>⑤ Hydrochlorothiazide</td></tr>
<tr><td>2023-4-35<br>[3-3]<br>Hard<br>(0/11)</td><td>Question: Which of the following statements about Sterile Water for Injection, as specified in the 12th edition of the Korean Pharmacopoeia, is correct?<br><br>① Microbial limit test is necessary.<br>② The endotoxin specification is the same as that of purified water.<br>③ The conductivity specification is the same as that of sterile purified water.<br>④ It is manufactured by placing purified water in containers and sterilizing.<br>⑤ It is manufactured by placing water for injection in containers and sealing.</td></tr>
<tr><td>2024-4-53<br>[3-4]<br>Medium<br>(6/11)</td><td>Question: Which of the following healthcare services is covered by the National Health Insurance of the Republic of Korea?<br><br>① Paternity test diagnosis<br>② Erectile dysfunction treatment<br>③ Aseptic preparation of injectable medications<br>④ Shingles (herpes zoster) vaccination<br>⑤ Medical treatment for improvement purposes</td></tr>
</table>

| Question | Translated Text |
|---|---|
| **2024-4-68**<br>[4-1]<br>Hard<br>(0/11) | Question: According to the "Pharmaceutical Affairs Act," which of the following is NOT required to be written on the container or packaging of a medication prepared by a pharmacist for the purpose of sale?<br><br>① Quantity prepared<br>② Date of preparation<br>③ Name of the preparer<br>④ Dosage and administration as written on the prescription<br>⑤ Name and location of the pharmacy where it was prepared |
| **2019-2-53**<br>[2-3]<br>Easy<br>(11/11) | The structural characteristics of procaine, used as a local anesthetic, were analyzed and confirmed using infrared spectroscopy and nuclear magnetic resonance spectroscopy.<br><Data (confidential)><br><br>Question: In which region (cm^-1) is the stretching vibration due to the amine group observed in the infrared spectrum?<br><br>① 850~700<br>② 1150~1000<br>③ 1750~1600<br>④ 2350~2200<br>⑤ 3550~3300 |
| **2023-2-27**<br>[2-2]<br>Hard<br>(1/11) | Question: What is drug A obtained from the following synthesis process?<br><br><Data (confidential)><br><br>① <Data (confidential)><br>② <Data (confidential)><br>③ <Data (confidential)><br>④ <Data (confidential)><br>⑤ <Data (confidential)> |

| Question | Translated Text |
|---|---|
| **2023-4-28**<br>[3-3]<br>Medium<br>(5/11) | Question: The following figure is a schematic diagram of the Common Technical Document (CTD). What content should be included in Module 4?<br><br><Data (confidential)><br><br>① Clinical data<br>② Quality data<br>③ Non-clinical data<br>④ Quality by Design (QbD) data<br>⑤ CMC (Chemistry, Manufacturing and Control) data |
