## Supplementary Figure 1 for "Proprietary and Open-Source Large Language Models on the Korean Pharmacist Licensing Examination: A Comparative Benchmarking Study"

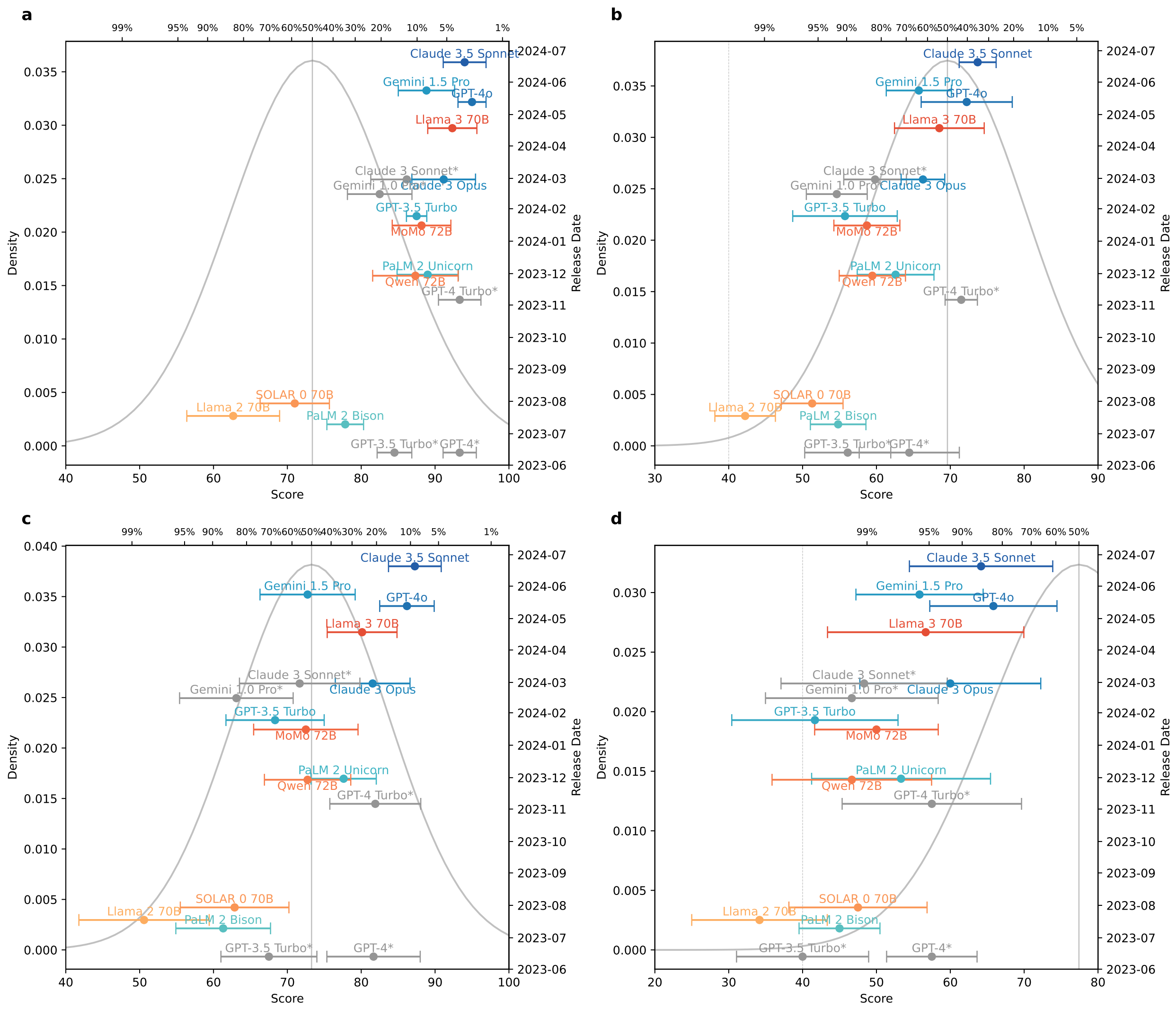


Figure 1: Temporal evolution of LLM performance by subject on the KPLE exam. (a–d) Performance trends for Biopharmacy (a), Industrial Pharmacy (b), Clinical & Practical Pharmacy (c), and Medical Health Legislation (d). Each point represents an individual model's raw score (bottom x-axis) and percentile rank (top x-axis) at the time of its release (y-axis). Human performance distributions are illustrated by the central gray curve. Passing thresholds are marked by vertical lines (60% overall, 40% per subject), and the 50th percentile reference line is included for context.
