## Supplementary Figure 2 for "Proprietary and Open-Source Large Language Models on the Korean Pharmacist Licensing Examination: A Comparative Benchmarking Study"

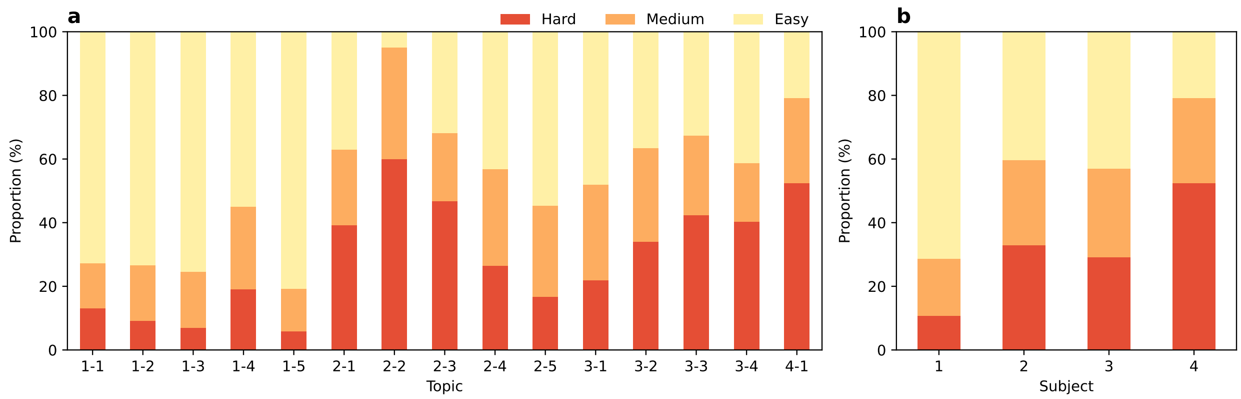


Figure 2: Item difficulty distribution for the low-scoring model group. (a) Proportion of items categorized as easy (light yellow), medium (orange), and hard (red) per individual topic for the low-scoring group. (b) Difficulty distribution by subject for the low-scoring group, following the same categorization. The overall pattern closely resembles that of the high-scoring group shown in Figure 5, demonstrating consistent difficulty trends across different model performance levels.
